# The Human Intolerome: a curated database to prioritize genomic variants in stillbirth, pregnancy loss, and neonatal death

**DOI:** 10.1101/2025.10.08.25336895

**Authors:** Svetlana A. Yatsenko, Amrita Nagasuri, Vishal Soman, Aryo Garakani, Mona Aminbeidokhti, Uma Chandran, Mahmoud Aarabi, Tomiko Oskotsky, Syed Hassan Bukhari, Catherine Hypes, Zhiping Gu, Ruth Monteiro, Sammi Smith, Bryan Walters, Michael P. Snyder, Christina G. Tise, Jonathan A. Bernstein, Gary M. Shaw, David K. Stevenson, Ruth B. Lathi, Marina Sirota, Aleksandar Rajkovic

## Abstract

**Background:** The application of next-generation sequencing in prenatal and neonatal genomic medicine provides definite diagnosis, impacts clinical decision-making and reproductive planning. Despite recent advances, interpretation of variants identified by genome/exome sequencing in cases lacking obvious phenotypic abnormalities (stillbirth, miscarriage, neonatal death) remains challenging.

**Methods:** To improve diagnostic accuracy, we created the Intolerome Database, a curated resource of 934 genes essential for viability in humans. This database has accumulated details on genes’ mechanisms of action, phenotypes, inheritance, the mortality timing, and supporting publications.

**Results:** The Intolerome includes 59 (6.3%) genes linked to the first/second trimester miscarriages, 525 (56.2%) genes associated with stillbirth/neonatal death, and 350 (37.5%) genes with variants that can cause lethality at any prenatal/postnatal stage. *De novo* inheritance was documented for 159 autosomal-dominant genes. Heterozygous potentially lethal variants in 39 autosomal-recessive genes were present in all ancestries in gnomADv4.1.0 at a frequency of ≥1/100 individuals, underlining the major pregnancy loss contributors.

**Conclusions:** The Intolerome serves as a comprehensive resource for identifying and interpreting genomic variants linked to fetal and neonatal mortality. It will support clinicians, laboratory professionals, and researchers in advancing the diagnosis and understanding of lethal genetic conditions, offering new insights into their clinical presentations and inheritance patterns.

## INTRODUCTION

Approximately 80% of all conceptions are lost and do not result in a live birth. Most of these losses occur during the early stages of embryo development from pre-implantation through the first four weeks of gestation.^1,2^ Miscarriage, defined as a spontaneous demise prior to fetal viability, is the most common adverse pregnancy outcome, documented in up to 30% of clinically recognized pregnancies.^1^ Approximately 50% of miscarriages that happen in the first trimester are due to *de novo* chromosomal anomalies, including aneuploidy, polyploidy, and less frequently unbalanced structural chromosome rearrangements.^1,3^ Up to 5% of women experience two or more pregnancy losses and are diagnosed with recurrent pregnancy loss (RPL). A significant portion of RPL cases involve euploid pregnancies^3,4^; however, establishing a molecular diagnosis is challenging, as the prenatal phenotype is often non-specific.

Pregnancies that proceed beyond 20 weeks of gestation have a much lower rate of aneuploidy and a higher chance of live birth. Stillbirth is defined as an intrauterine death of a baby after 20 weeks of gestation. In developed countries, the incidence of stillbirth ranges between 1.7 and 6 per 1,000 births. In the United States, a rate of 5.5-5.8 stillbirths per 1,000 livebirths^5^ is nearly similar to a neonatal mortality rate at 5.8 per 1,000 livebirths.^6,7^ Globally, nearly 2-3 million stillbirths occur annually.^8^ After the exclusion of chromosomal causes, obstetric complications, infections, fetal congenital abnormalities, placental and metabolic pathologies, specific causes of euploid pregnancy loss, stillbirth and neonatal death cannot be explained in more than 60% of cases.^9^ Pedigree analysis of couples with recurrent miscarriages reveals an increased co-occurrence of adverse reproductive outcomes including miscarriages at various developmental stages, intrauterine growth restrictions, stillbirth, preterm birth, and neonatal mortality, suggestive of intertwined genetic etiologies.^10,11^

The implementation of exome and genome sequencing has significantly improved diagnosis for children with congenital anomalies, metabolic, and neurodevelopmental conditions. The diagnostic yield of exome (ES) and genome sequencing (GS) in the pediatric population ranges from ∼20% to 40%, depending on the specific clinical manifestations.^12–15^ Despite the knowledge that prenatal lethality may be caused by genetic conditions, the utilization of next generation sequencing (NGS) technologies is hampered by indeterminate or atypical fetal phenotype, providing a diagnosis in 6%-10% of unexplained euploid RPL and stillbirth.^16,17^ A few NGS studies have focused on small cohorts of late gestation cases with congenital abnormalities which were used for a phenotype-driven variant prioritization in genes associated with known syndromic conditions.^18,19^ In cases of early miscarriage and death of a baby with no apparent problems, finding genetic causes in the absence of fetal and placental pathologies is challenging and requires an agnostic approach to ES and GS analysis.

Information about genomic variants in specific genes labeled as “lethal genes”, that may result in stillbirth or a neonatal death, their association with potentially a more severe *in utero* phenotype, incompatible with life, the susceptibility to modifying factors and environmental exposures are scattered in the literature and have not been thoroughly studied. To facilitate systematic analysis and improve our understanding about causes of prenatal and neonatal lethality, we developed the Human “Intolerome” Database (https://rpldb.org/intolerome), comprising a set of genes, variants in which are not tolerated, and are associated with embryonic lethality, spontaneous intrauterine fetal demise, stillbirth, and neonatal mortality. This online resource has aggregated information about genes and chromosomal loci, prenatal phenotypes, and perinatal outcomes from published reports of genetic aberrations found in human samples.

## METHODS

### Literature search strategy

PubMed, Google Scholar, and the Online Mendelian Inheritance in Man (OMIM) databases were searched up to December 2023 for original and review articles on cohorts and individual cases, reporting genetic findings in patients with spontaneous miscarriage of euploid conceptions, RPL, stillbirth, neonatal death, and monogenic causes of infantile mortality. The keywords used to retrieve articles of interest are given in the Supplemental Table S1. The literature references published in English were collected and manually curated to extract the following parameters: gene name; genomic variant; sex of the fetus or product of conception; prenatal ultrasound findings; postnatal manifestations; obstetric complications; associated disease; variant effect on a fetus, mother or both, contributing to lethality; mode of inheritance, reproductive outcomes; lethality timepoint; family history and genetic testing of family members. Sequencing findings in miscarriage samples positive for a chromosomal aneuploidy, hydatidiform mole or triploidy were excluded, unless genomic variants involved genes responsible for chromosome segregation.

### Annotations of genes and variants

Based on the available information from the original manuscripts, genes associated with lethality were classified into three categories: 1) “known lethal genes” with five or more independent studies and a well-established gene-lethality association; 2) “emerging lethal genes” with lethality evidence reported in 2-4 articles; and 3) “candidate genes” with a single publication reporting a lethal outcome. Variants found in each gene were categorized based on the molecular type (single nucleotide variant (SNV) or structural variant (SV)) and, when possible, based on the predicted functional impact (a loss-of-function, gain-of-function). For each gene, available OMIM phenotypes were analyzed for evidence of prenatal and neonatal lethality, inheritance mode and penetrance. Clinical manifestations were reviewed and mapped to affected physiological systems. Genes associated with multiple congenital anomalies involving three or more systems were coded as multiple (≥3). The timing of lethality was assigned to first-, second-, or third-trimester and/or postnatal periods based on clinical data and pedigree analyses reported in the source manuscripts. Dosage sensitivity (haploinsufficiency and triplosensitivity) and genomic coordinates were annotated for each gene using The National Center for Biotechnology Information database (https://www.ncbi.nlm.nih.gov). Pathway enrichment analysis was performed using the PANTHER (Protein Analysis Through Evolutionary Relationships) Gene Ontology database https://pantherdb.org/ for *Homo Sapiens* model organism.

### Estimation of “lethal” allele frequencies and carrier frequency in human populations

To estimate the carrier frequency for “lethal” variants in the genes associated with autosomal recessive conditions (AR) included in the Intolerome database, we calculated gene carrier frequency (GCF) as the cumulative frequency of pathogenic and likely pathogenic variants as well as predicted lethal variants. The predicted lethal variant is defined as a protein-coding variant that has never been observed in a homozygous state, but would be expected to be present based on the heterozygous carrier frequency. Due to the large number of participants from various ancestries, we utilized Genome Aggregation Database (gnomAD) v4.1.0 encompassing ∼731,000 exomes for GCF estimation. Variants with *nhomalt = 0* (i.e., no observed homozygous alternate genotypes) counts in exomes were extracted from gnomAD v4.1.0 and cross-checked for *nhomalt = 0* counts in ∼76,000 genomes. For each variant, Allele Count (AC), Allele Number (AN) and Allele Frequency (AF) were extracted and stored in chromosome-sorted TSV files. Prior to frequency calculations, variants with low quality or a problematic flag, including low complexity regions, counts from the non-coding transcripts, variants overlapping segmental duplication regions and pseudogenes^20^ were filtered out. We also excluded variants falling in the 5’ UTR, 3’ UTR, intronic regions, as well as synonymous variants. These data were then used to estimate variant carrier frequency (VCF) and gene carrier frequency (GCF), as described previously^21^ and modified for Hom=0:

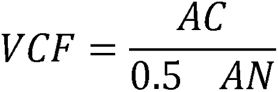

VCF is defined as the proportion of individuals in a population that carry the variant of interest. AC represents the number of alternate alleles, and AN indicates the total number of alleles.

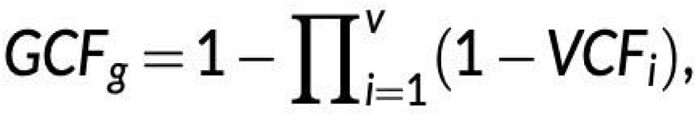

GCF is defined as the proportion of individuals in a population heterozygous for all P/LP and lethal variants in a given gene. VCF values were used to calculate GCF for variants in the filtered Intolerome gene list. VCF_i_ is the VCF for variant *i* of gene *g,* and *v* is the number of variants in a gene.

Carrier frequency data for VCF and GCF across chromosomes were concatenated into TSV files respectively. Using GCF calculations, the expected homozygote probability and count were estimated. Chi-square tests were used to identify genes with statistically significant number of variants with the observed zero homozygotes against the expected value of, and chi-square tests were performed against the observed value of zero homozygotes. The resulting gene list from GCF calculations and chi-square testing was further filtered to include only the Intolerome-related autosomal recessive (AR) genes with p-values < 0.05. All gnomAD VCF queries and carrier frequency computations were executed in Python (v3.11.9) and Bash (v4.4.19).

## RESULTS

Comprehensive literature search yielded 889 publications describing monogenic causes of euploid pregnancy loss, including variants in maternal genomes conferring genetic predisposition to miscarriage and variants in the conceptuses that impact their viability or decrease survival. A total of 934 genes were identified, including 582 (62.3%) “known lethal genes”, 270 (28.9%) “emerging lethal genes”, and 82 (8.8%) “candidate genes” for human lethality. Although previous studies indicated the presence of 624 known lethal genes^22^ in the OMIM database, our search revealed only 582 genes with sufficient evidence to be categorize as the established lethal genes. Other genes were included in the “emerging” or “candidate” gene categories. The Human Intolerome gene database (https://rpldb.org/intolerome) is searchable with hyperlinks to the relevant phenotype entries in the OMIM database and to the PubMed identifier numbers.

### Maternal genetic factors predisposing to pregnancy loss

Deleterious variants in 16 genes (16/934, ∼2%) have been associated with increased susceptibility to miscarriage and abnormal gametogenesis including recurrent aneuploidy, abnormal placental development and embryonic arrest (Figure 1A). The genetic predisposition to RPL, particularly in the first trimester, has been linked to maternal hypercoagulability conditions such as thrombophilia. Pathogenic variants in the prothrombin (*F2*, MIM*176930 (HGNC:3535)) and the coagulation factor V (*F5,* MIM*612309 (HGNC:3542)) gene confer a risk for placental thrombosis, inflammation, and endothelial dysfunction that have been observed at the maternal-fetal interface in women with RPL.^23^

**Figure 1.**
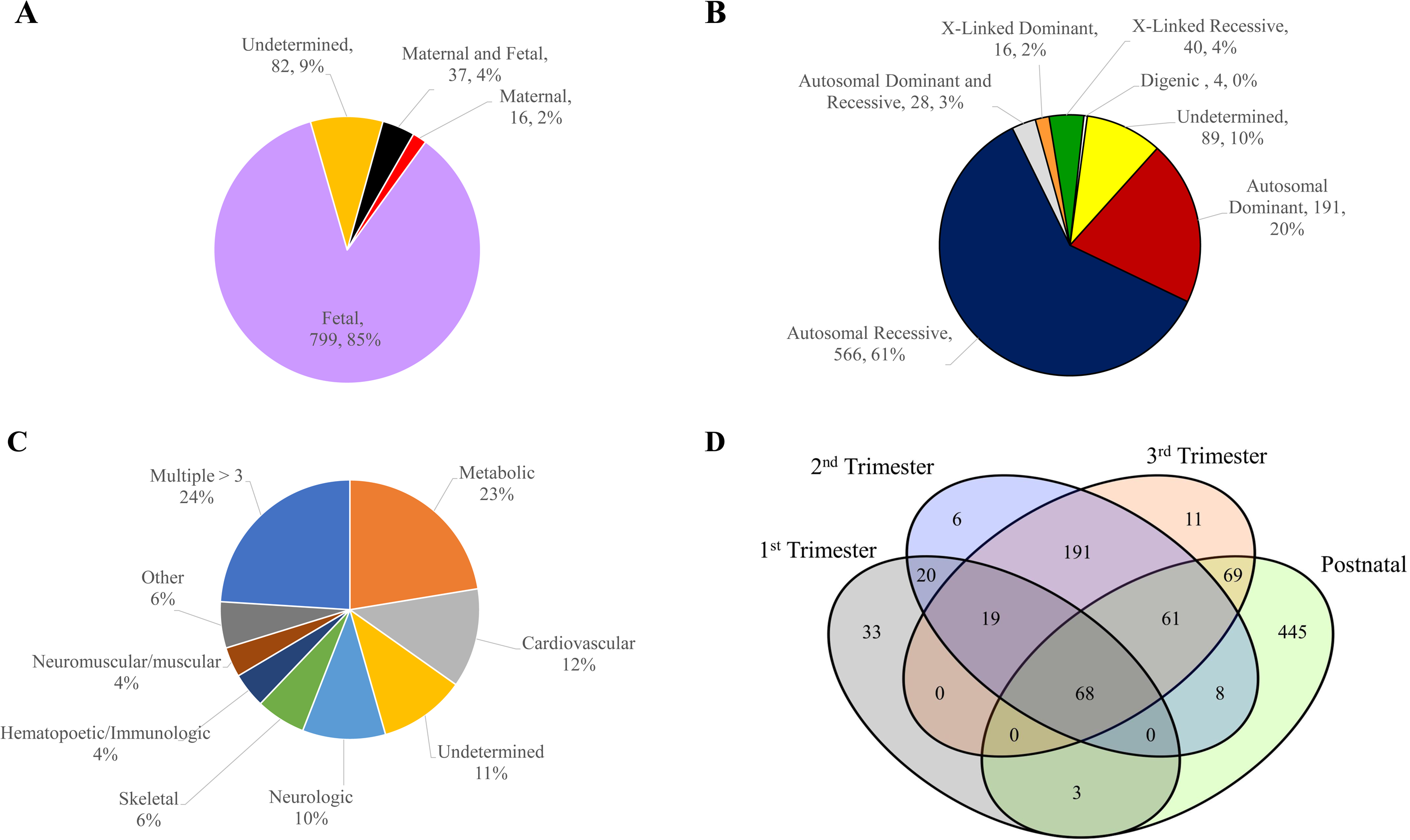
Characteristics of human genes with lethal phenotypes. **(A)** Distribution of maternal or fetal genes contributing to prenatal and perinatal lethality. **(B)** Mode of inheritance. (**C**) Distribution of genes based on their clinical manifestations and the major organ systems involvement. (**D**) The lethality timeline and the number of genes associated with the first, second, or third trimester pregnancy losses or a death during postnatal period.

Maternal variants in genes (e.g., *NLRP7* MIM*609661(HGNC:22947)*, KHDC3L* MIM*611687(HGNC:33699), *MEI1* MIM*608797 (HGNC:28613), *TOP6BL* MIM*616109 (HGNC:26197), *REC114* MIM*618421 (HGNC:25065)) controlling meiotic double-strand break formation, chromosomal segregation, and the oocyte genome epigenetic reprogramming have been identified in women with recurrent complete hydatidiform moles and miscarriages.^24^ Growing evidence suggests that metabolic factors, including glucose and lipid metabolism, may play critical roles in physiological maternal adaptation to pregnancy and in the pathophysiology of unexplained RPL,^25,26^ while maternal-fetal synergy is essential to ensure efficient nutrient transfer, primarily via the placenta, and proper fetal development. Our search identified 37 genes (37/934, 4%) (Figure 1A), in which maternal and fetal variants are associated with metabolic and immunological conditions, affect cellular homeostasis, and/or result in oxidative stress and inflammation. An additional 82 genes (82/934, ∼9%) were implicated in RPL, however the etiologic contribution – maternal, fetal, or combined– has not been yet established. These genetic predispositions may be substantially modified by environmental factors and toxic exposures that perturb maternal–fetal molecular signaling during early gestation.

### Mendelian conditions associated with miscarriage and fetal lethality

Single-gene Mendelian conditions are important and understudied contributors to miscarriages and stillbirth. The “Intolerome” database comprises 799 genes linked to prenatal and perinatal lethality. Autosomal dominant conditions were documented for 191 (20%) genes, including 159 genes with *de novo* heterozygous variants in miscarriage tissues and/or affected neonates, and 32 genes with variants transmitted from a parent. X-linked lethal disorders were recognized for 56 (6%) genes. Homozygous and compound heterozygous variants in 566 (61%) autosomal recessive (AR) genes were the major cause of neonatal death and perinatal loss of euploid conceptions described in literature (Figure 1B). These variants can be inherited from healthy parents, carriers of heterozygous variants leading to recurrent miscarriages, stillbirths and infantile deaths. Our search also identified 89 (10%) genes (Figure 1B) with inconsistent inheritance reported among RPL families. It is unclear if maternal heterozygosity for a specific variant predisposes to miscarriage of a fetus with an inherited heterozygous variant or the viability of conceptions with biallelic variants is compromised. Lastly, a small portion of genes (28, 3%) was reported to manifest with lethal phenotypes in carries of heterozygous and biallelic variants in the same gene or due to variants in multiple genes (4 genes with digenic inheritance).

### Genes associated with autosomal dominant conditions

Genomic alterations in 191 genes linked to autosomal dominant (AD) lethal conditions are less frequently inherited from unaffected parents. Many of these AD disorders can be diagnosed in utero due to an associated abnormal phenotype affecting the cardiovascular (74 AD genes, 39%), brain (21 AD genes, 11%), skeletal (14 AD genes, 7%), or multiple systems (50 AD genes, 26%). *De novo* pathogenic variants in 159 of these genes have been documented in sporadic cases, although multiple affected conceptions were also reported. Both incomplete penetrance and parental germline mosaicism have been reported in literature explaining the reoccurrence and transmission of a lethal allele from seemingly asymptomatic parents.^27,28^ Maternal carrier status for some AD conditions, such as the Long QT syndrome, may also poses an additional risk of adverse pregnancy outcomes.^29^ At least 93 (93/191, 48.7%) of these AD genes are dosage sensitive with lethal phenotypes caused by haploinsufficiency. Multiple genomic disorders^30^ and microdeletion syndromes comprise dosage sensitive genes, that are associated with high risk of mortality, particularly for conditions with cardiovascular developmental defects. Multiple studies utilizing chromosomal microarray analysis in spontaneous miscarriages have discovered recurrent microdeletions in the 7q36.1, 8p23.1, and 22q11.21 regions comprising dosage sensitive genes *KCNH2* (HGNC:6251), *GATA4* (HGNC:4173), and *TBX1* (HGNC:11592), respectively.^31–34^

### Genes associated with autosomal recessive conditions

In miscarriages and cases of a neonatal death, biallelic variants were documented for 566 genes, including 425 (75.1%) genes implicated in the well-established AR conditions, 121 (21.4%) genes with emerging evidence, and 20 (3.5%) candidate genes. In addition, 32 genes were reported with both AD and AR, or digenic inheritance. Based on current literature, variants in at least 25% of these genes are associated with malformations involving multiple systems. The ACMG recommended preconception carrier screening panel for 112 pediatric moderate and severe conditions comprises 45 (45/566, 7.9%) of these AR lethal genes.^35^ Only 88 genes out of all AR genes in the Intolerome database (14.7%) are present in the proposed 286 pan-ethnic carrier screening gene list, proposed based on the data form gnomAD v4.1.0.^36^ Both preconception carrier screening panels were structured based on the carrier frequency of pathogenic/likely pathogenic (P/LP) variants in the general human population. Lethal genomic variants are less likely being classified as P/LP if the resulting phenotype is an early *in utero* death when anatomical or functional anomalies cannot be detected. By analysis of variants that are very frequent in humans but have never been documented in a homozygous state in gnomAD v4.1.0 exomes or genomes (Figure 2), we identified top 39 genes with potentially lethal variants, yet many of them are not classified as P/LP (Table 1, Supplemental Table S2). Based on the carrier frequencies for these genes from 1/10 to 1/100 in various genetic ancestry groups, up to 12% of non-viable conceptions might be due to biallelic variants in one of these genes. Our previous studies also demonstrated high frequency of heterozygous, loss-of-function variants in 11 out of these 39 genes (*ACE* (HGNC:2707), *CEP290* (HGNC:29021), *COL7A1* (HGNC:2214), *MED16* (HGNC:175560), *NUP214* (HGNC:8064), *PKHD1* (HGNC:9016), *PRKRA* (HGNC:9438), *RYR1* (HGNC:10483), *SPEG* (HGNC:169010), *SYNE1* (HGNC:17089), *WNK1*(HGNC:14540)).^37^

**Figure 2.**
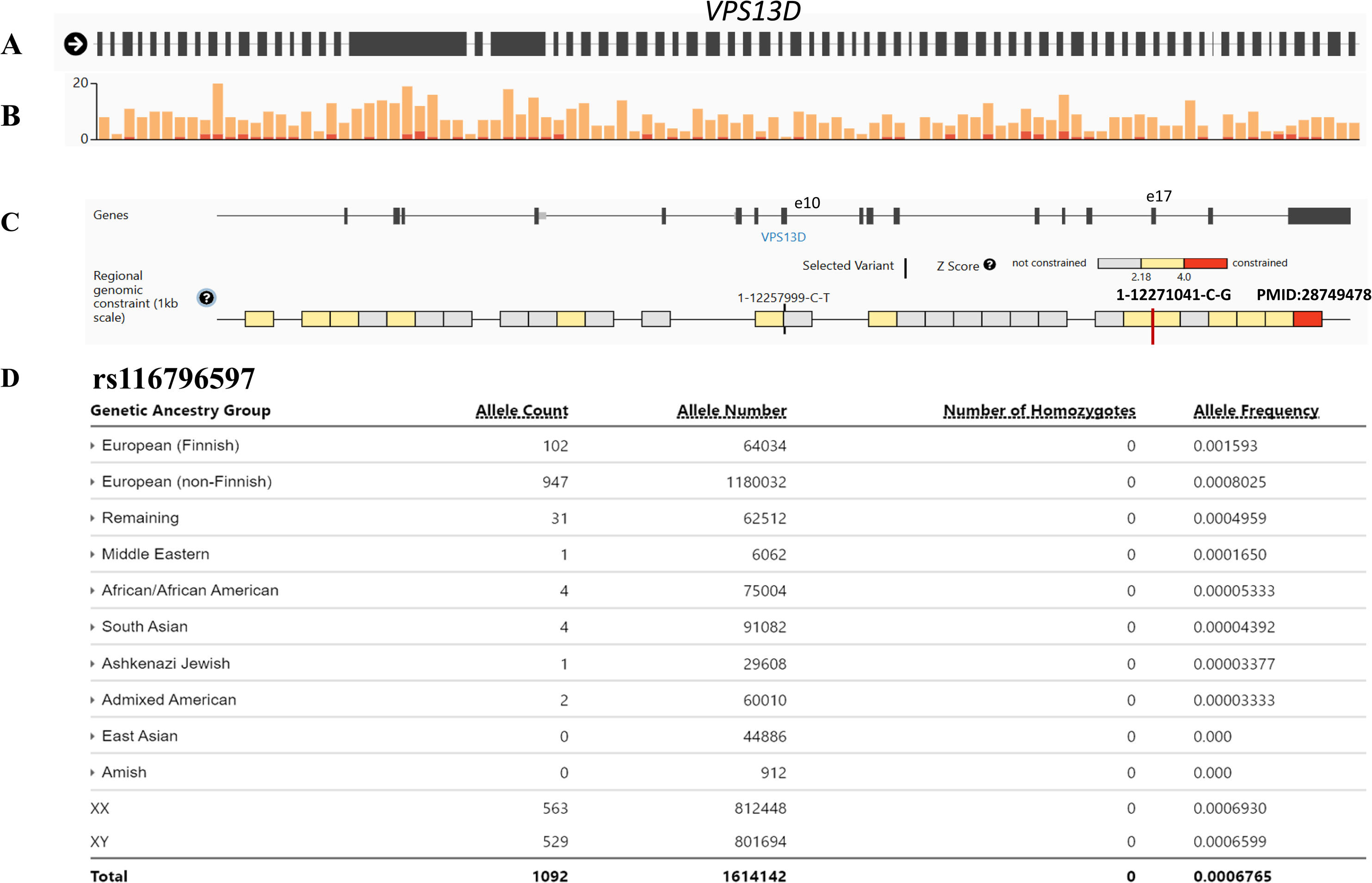
Identification of a potentially lethal variant in the *VPS13D* gene. Screenshots of the gnomAD browser for the *VPS13D* gene are shown to exemplify characteristics of potentially lethal variants. **(A)** Genomic structure of the *VPS13D* gene, comprising 69 exons. The human *VPS13D* gene spans approximately 282 kb. **(B**) Nearly 13,200 variants are documented in the database, including ∼5400 variants with no homozygous allele counts. Pathogenetic and likely pathogenic alleles are displayed in red, variants of uncertain significance are indicated in orange. **(C)** A potentially lethal variant p.Arg336Cys (1-12257999-C-T; rs116796597) is located in exon 10. This region of genome has been overall depleted for variations with the assigned Z constraint score of 3.27 (yellow blocks), indicating that the number of observed variants is fewer than expected. A homozygous variant p.Arg674Gly (1-12271041-C-G, rs756025227, marked by red vertical line) in the *VPS13D* gene, reported in a case of intrauterine fetal demise at 22 weeks of gestation (PMID:28749478), is also located in the constraint region of exon 17. (**D**) Frequency of the rs116796597 allele in genetic ancestry groups. Despite the significant rate of heterozygous carriers for this variant, no homozygous alleles were identified (p-value = 0.0014), suggestive of lethality in homozygotes.

**Table 1.** Genes with P/LP and potentially “lethal” variants with the carrier rate ≥1/100 in the general human population.

| Gene | OMIM gene | Gene category | Disease | OMIM Phenotype | GCF HMZ-Max | GCF HMZ-Min | Population with max GCF |
| --- | --- | --- | --- | --- | --- | --- | --- |
| *ACE* | 106180 | Known | Renal tubular dysgenesis | 267430 | 0.111222 | 0.026486 | EAS |
| *CEP290* | 610142 | Known | Meckel syndrome | 611134 | 0.089617 | 0.023860 | AFR |
| *COL7A1* | 120120 | Candidate | Epidermolysis bullosa | 226600 | 0.103893 | 0.032703 | AFR |
| *DCHS1* | 603057 | Emerging | Van Maldergem syndrome | 601390 | 0.101206 | 0.035320 | AFR |
| *DNAH11* | 603339 | Emerging | Ciliary dyskinesia with or without situs inversus | 611884 | 0.184753 | 0.034235 | AFR |
| *DNAH5* | 603335 | Known | Primary ciliary dyskinesia (PCD) | 608644 | 0.184576 | 0.053252 | AFR |
| *DNAH9* | 603330 | Emerging | Primary ciliary dyskinesia | 618300 | 0.160313 | 0.061420 | EAS |
| *DNHD1* | 617277 | Emerging | Spermatogenic failure, congenital heart defects | 619712 | 0.159237 | 0.039788 | AFR |
| *DNM2* | 602378 | Known | Lethal congenital contracture syndrome | 615368 | 0.188203 | 0.028584 | AMR |
| *DYNC2H1* | 603297 | Known | Short-rib thoracic dysplasia | 613091 | 0.134200 | 0.027910 | AFR |
| *EPG5* | 615068 | Known | Vici syndrome | 242840 | 0.088853 | 0.024196 | AFR |
| *ERBB3* | 190151 | Known | Lethal congenital contractural syndrome | 607598 | 0.318752 | 0.018291 | NFE |
| *FOXRED1* | 613622 | Known | Mitochondrial complex I deficiency | 252010 | 0.301466 | 0.045612 | FIN |
| *FRAS1* | 607830 | Known | Fraser syndrome | 219000 | 0.143374 | 0.041091 | AFR |
| *FREM2* | 608945 | Emerging | Cryptophthalmos | 123570 | 0.126521 | 0.028508 | AFR |
| *HSPG2* | 142461 | Known | Dyssegmental dysplasia | 224410 | 0.391109 | 0.107049 | MID |
| *IFT140* | 614620 | Emerging | Jeune Asphyxiating Thoracic Dystrophy | 266920 | 0.087139 | 0.016812 | MID |
| *KCNH2* | 152427 | Known | Long and short-QT-syndrome; Brugada syndrome | 613688; 609620 | 0.076943 | 0.029829 | EAS |
| *LAMA2* | 156225 | Emerging | Dilated cardiomyopathy | 612877 | 0.135124 | 0.029082 | AFR |
| *LAMA3* | 600805 | Known | Epidermolysis bullosa, junctional, severe | 226700 | 0.116933 | 0.050663 | AFR |
| *MPDZ* | 603785 | Emerging | Hydrocephalus, with or without brain or eye anomalies | 615219 | 0.106391 | 0.015608 | AFR |
| *MYH6* | 160710 | Emerging | Sudden infant death syndrome | 272120 | 0.088136 | 0.026388 | AFR |
| *MYO18B* | 607295 | Known | Klippel-Feil syndrome | 616549 | 0.109325 | 0.017497 | AFR |
| *NEB* | 161650 | Known | Nemaline myopathy | 256030 | 0.235256 | 0.102619 | AFR |
| *NUP214* | 114350 | Candidate | Encephalopathy, acute, infection-induced | 618426 | 0.120942 | 0.032428 | AFR |
| *OBSCN* | 608616 | Known | Dilated cardiomyopathy | 612877 | 0.404268 | 0.158945 | AFR |
| *PIEZO1* | 611184 | Known | Lymphatic malformation 6 | 616843 | 0.193900 | 0.052300 | AFR |
| *PIP5K1C* | 606102 | Known | Lethal congenital contractural syndrome | 611369 | 0.045237 | 0.019256 | AMR |
| *PKD1L1* | 609721 | Known | Heterotaxy, visceral | 617205 | 0.128115 | 0.029411 | AFR |
| *PKHD1* | 606702 | Known | Polycystic kidney disease | 263200 | 0.130747 | 0.059672 | AFR |
| *PLEC* | 601282 | Known | Epidermolysis bullosa simplex with muscular dystrophy | 226670 | 0.272249 | 0.097242 | AFR |
| *PRKRA* | 603424 | Candidate | Dystonia | 612067 | 0.395031 | 0.151260 | NFE |
| *RIPK4* | 605706 | Known | Popliteal pterygium syndrome | 263650 | 0.077609 | 0.027713 | MID |
| *RYR1* | 180901 | Known | Neuromuscular disease, congenital | 255320 | 0.136919 | 0.054407 | AFR |
| *SPEG* | 615950 | Known | Centronuclear myopathy | 615959 | 0.132510 | 0.048472 | AMR |
| *SYNE1* | 608441 | Candidate | Arthrogryposis | 618484 | 0.303943 | 0.071069 | AFR |
| *VPS13D* | 608877 | Emerging | Spinocerebellar ataxia | 607317 | 0.090782 | 0.050231 | EAS |
| *WDR81* | 614218 | Emerging | Hydrocephalus, congenital with brain anomalies | 617967 | 0.072894 | 0.023514 | MID |
| *WNK1* | 605232 | Candidate | Neuropathy, hereditary sensory and autonomic | 201300 | 0.115628 | 0.044672 | EAS |

Ethnic groups: African/African American (AFR), Latino/Admixed American (AMR), East Asian (EAS), Finnish (FIN), Non-Finnish European (NFE).

### Genes associated with X-linked conditions

To date, the Intolerome database contains 56 X-linked genes, including 40 genes that are expected to affect males only (X-linked recessive) and 16 genes reported to affect both males and females (X-linked dominant), although in regards to lethality both XX and XY conceptions might be at risk. The majority of genes (47) are well established, while 9 genes are emerged from the recent studies.

### Affected organ systems

Based on the clinical manifestations in prenatal and postnatal cases, 833 genes were characterized according to the body systems involved. For the remaining 101 genes, the major affected organ system could not be assigned due to either early timing of a pregnancy loss or lack of any obvious anomalies. The highest number of genes in the database is associated with severe disorders characterized by multisystem malformations (3 or more affected organ systems), followed by metabolic conditions and cardiovascular developmental problems. Other frequent presentations included brain anomalies, skeletal defects, isolated hematologic and neuromuscular phenotypes (Figure 1C).

### Lethality timepoint

For each gene, we reviewed clinical reports and available pedigrees to extract information about gestational age of the fetus at the time of miscarriage or a childhood death. Lethality timepoints (the first, second, third trimester, or postnatal period) were assigned according to the gestational age at which the death occurred (5-12 weeks, 13-24 weeks, 25 weeks – birth, or after birth, respectively. For multiple genes lethality was documented across multiple timepoints (Figure 1D). For the majority of genes, deaths were observed during the third trimester and the postnatal period. Only 59 genes were associated with miscarriages exclusively in the first and second trimesters. Although the number of genes associated with early miscarriages is small, we attempted to identify the primary functions and the molecular pathways annotated with 59 genes linked to the first and second trimester and compare to the pathways associated with 525 genes linked to stillbirth and postnatal death (Figure 3). Interestingly, in the early embryo development, interaction of individual cells and cell homeostasis appears to be a fundamental difference from the later stages. The integrin signaling pathway facilitates cell-to-cell communication as wells as promotes cell adhesion, migration, and proliferation. The integrin and angiogenesis pathways are critical for tissue development, survival, and differentiation (Figure 3A). In the third trimester and the postnatal period, the key pathways are implicated in tissue/organ metabolism and energy regulation (Figure 3B).

**Figure 3.**
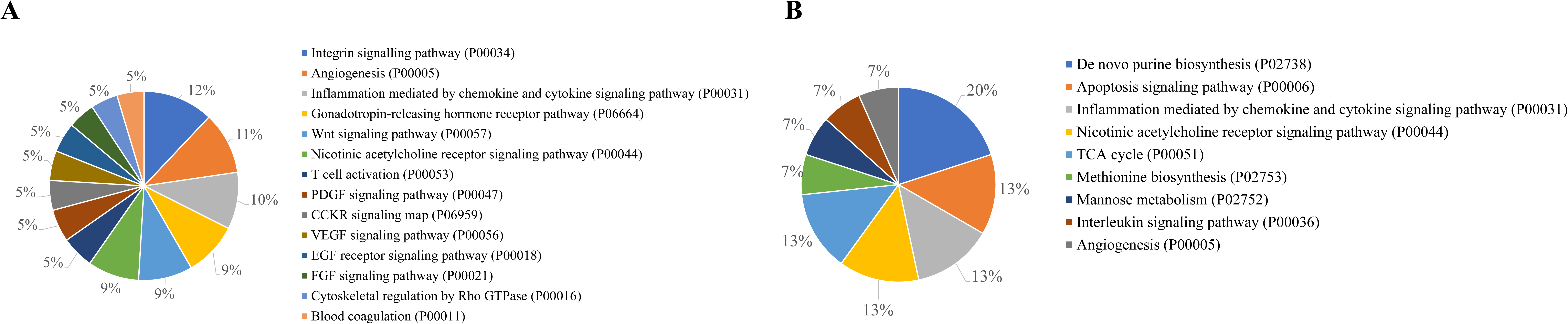
Top Pathways across prenatal and neonatal stages. **(A)** Pathways identified in genes associated with first and second trimester pregnancy loss. **(B)** Pathway enrichment analysis in genes linked to stillbirth and postnatal death.

## DISCUSSION

Thousands of genes are essential for embryo development and growth, yet the genetic etiology underlying miscarriage of chromosomally normal conceptions remains largely underdetermined. With the application of next-generation sequencing to the diagnosis of RPL and stillbirths, numerous genetic changes can be identified in the fetus and the parents. However, interpreting the clinical significance of these variants and definitively attributing them as the cause of a pregnancy loss can be challenging, especially if the variants have not been linked to a known disease or if a gene has not been reported in miscarriages.

The Human Intolerome Database represents a resource of human genes, variants in which were observed in fetuses and neonates associated with lethal outcomes. This is a curated and dynamically updated list of genes that can be used for prioritization of genomic variants in pregnancy loss, stillbirth, and neonatal death. It is particularly powerful in cases lacking obvious anatomical or physiological abnormalities, where the phenotype-driven approach for interpretation of exome and genome data is a major limitation. This Intolerome gene – driven evaluation can also be applied for cases with structural fetal anomalies, when the initial analysis is nondiagnostic. Even for a well-characterized condition, the phenotype assessed in utero by imaging is limited due to technological and biological constraints. Moreover, for ∼70% of all genes the disease phenotypes are unknown or have more severe in utero manifestations incompatible with life. A targeted approach has the potential to yield specific diagnosis after a detailed examination of relevant genomic loci in cases with an atypical presentation.

Interpretation of variants’ pathogenicity and their classification in clinical decision making requires knowledge of the patient’s phenotype, mode of inheritance, functional mechanism, and the frequency of a variant in the general population. The gnomAD database v4.1.0 has accumulated a comprehensive catalogue of more than 909 million variants identified in 807,162 individuals through exome and genome sequencing initiatives. Rarity of a variant is considered as a prerequisite for pathogenicity for a disease in liveborn individuals. However, in cases of AR fatal conditions, a combination of two lethal alleles is not likely to appear among living individuals, but the overall heterozygote carrier frequency of lethal variants is not necessarily low in the reference population. We used this fact to mine for potentially lethal variants that are never observed in a homozygous state despite a significantly higher number of the expected homozygotes. Among the 566 AR genes documented in the Intolerome database, 39 genes had an extended number of heterozygous protein-coding but unclassified variants, giving an overall gene carrier frequency from 1/10 to 1/100 in various genetic ancestry groups. Interestingly, potentially lethal variants in some genes are much more prevalent in the specific ancestries than other ethnicities, which might be explained be an increased load of the deleterious genetic changes and founder effect, contributing to lethality.

A traditional trio-based approach (parents and conception) to diagnosis of pregnancy loss is not always possible due to early embryo demise or inability of obtaining fetal samples for genetic testing. In the absence of a proband’s sample, analysis of genes included in the Intolerome database provides an alternative solution for data interpretation of the parental samples. For the parents whose genetic analyses revealed variants of uncertain significance in the same “lethal” gene, these variants can be prioritized for further study, overcoming the bottleneck of investigating thousands of potential variants.

Currently, the number of genes implicated in the first trimester miscarriages is low, although our analysis identified a few molecular pathways that are critical for the early stages of human development. Analyzing genes within the same disrupted pathways can lead to the discovery of human-specific genetic causes of lethality, even if those genes cannot be studied through model organisms. By focusing on genes compiled in the Intolerome database, researchers and clinicians have an opportunity to discover and study novel disease-causing and “lethal” variants, and further advance the field of reproductive genetics by increasing the number of genomic changes that are known to be intolerant to variation in humans. It may also helpful for the identification of non-genetic modifiers, such as environmental and toxic exposures, affecting different developmental pathways during the first trimester of pregnancy when an embryo is most vulnerable to birth defects and lethality.

In summary, the Human Intolerome Database is the first resource of systematically evaluated known and candidate genes associated with prenatal and perinatal lethality. It will improve clinical diagnosis, facilitate identification of novel candidate genes, and inform reproductive planning. Identification of genes and variants that are causing or predisposing to pregnancy loss has direct implications for a woman’s reproductive and general health. Improved diagnosis also decreases the risk of transmitting pathogenic variants through assisted reproductive technologies and is critical for developing future successful treatment approaches. The Intolerome gene list can also be used for designing a preconception carrier screening panels to identify couples at-risk. By leveraging existing knowledge, future studies can identify molecular pathways, lethality-associated genetic variants, differentially expressed genes, and environmental contributors to prenatal and perinatal loss.

## Data Availability

The data underlying this article are available in the Human Intolerome Database (https://rpldb.org/intolerome). Additional data will be shared upon reasonable request to the corresponding author.

## Acknowledgments Funding Statement

This study was supported by the National Institutes of Health (NIH) (Grant number: R01 HD105256).

## Author Contributions

Conceptualization: S.A.Y., A.R.; Data curation: A.N., A.G., M.A., S.A.Y.; Formal analysis: A.N., V.S., A.G., S.H.B., M.A., S.A.Y; Funding acquisition: R.B.L., M.P.S., M.S., A.R.; Investigation: A.N., A.G., V.S., M.A., S.H.B., T.O., C.G.T., J.B., M.S., S.A.Y.; Methodology: V.S., U.C., M.A., M.A., A.R., S.A.Y.; Supervision: U.C., M.A., C.H., M.S., A.R., S.A.Y.; Visualization: C.H., Z.G., R.M., S.S., B.W.; Writing-original draft; M.A., M.A., A.N., V.S., C.G.T., J.B., G.M.S., D.K.S., R.B.L., M.P.S., M.S., A.R., S.A.Y.; Writing-review & editing: all authors.

## Declaration

This article included secondary analysis of published data. No human or animal participants were involved in this study.

